# Radio-frequency ablation versus hepatic surgery for the treatment of hepatocellular carcinoma: a systematic review and meta-analysis

**DOI:** 10.1101/2023.04.04.23288143

**Authors:** Avichal Dani, Khushi Vishal Gandhi, Dev Desai

**Affiliations:** Smt. NHLMMC, Ahmedabad, India

## Abstract

**Background:** Hepatocellular carcinoma (HCC) is the most common malignant tumor composed of cells resembling hepatocytes. It is the fourth most common cause of cancer□related death on earth. Treatment involves radio frequency ablation (RFA)or hepatic resection (HR). This is a review & evaluation of evidence comparing either methods by using meta-analysis technique.

**Materials and methods:** We conducted a database search of the PUBMED, GOOGLE SCHOLAR, Cochrane, EMBASE etc. in which total of 36 observational studies and 3 RCTs following PRISMA guidelines till September 2020 and matching inclusion and exclusion criteria were collected. These studies include total 16,700 patients out of which 8565 were treated with RFA & 8135 with surgery. The following search strings were used: “ RFA vs HR”, “hepatocellular carcinoma treatment “. The primary end point was overall survival rate in 3&5 years respectively, including hospital stay duration & local recurrence. RevMan 5.3 was used for appropriate statistical tests. Fixed and Random Effect Model Tests was used and p<0.05 was considered statistically significant.

**Results:** Meta-analysis showed that RFA was associated with significant decrease in the length of hospital stay for RCTs (SMD = -2.171, CI = -2.381 to - 1.962, p=<0.001) and non-RCTs (SMD = -1.048, CI = 1.492 to -0.937, p=<0.001) respectively. However, it was also associated with significant increase incidence of recurrence (RR = 1.749, 95% CI = 1.444 to 2.119, p=<0.001) and significantly poorer 3-year (RR = 0.850, 95% CI = 0.772 to 0.935, p=0.001); (RR = 0.941, 95%CI = 0.927 to 0.956, p=<0.001) survival chances for RCTs and non-RCTs respectively. 5-year survivability was (RR=0.856, 95% CI = 0.835 to 0.878, p=0.001).

**Conclusion:** Although RFA was associated with decreased duration of hospital stay, it was associated with increased chances of recurrence compared to hepatic resection. 3-year survival rate was also poorer.

## Introduction

Primary liver carcinoma is a prevalent cancer with a high fatality rate. Primary liver cancer has become more common in recent years, causing significant worry. (1) With an estimated 500,000 fatalities each year (2), hepatocellular carcinoma (HCC) is the world’s fifth most frequent malignancy. Due to the absence of identifiable symptoms in the early stages of primary liver cancer, most cases are detected in the middle to late stages. Surgery, chemotherapy, radiation, and biotherapy are all popular therapies for liver cancer. (3) (4) Advances in diagnostic imaging and the widespread use of screening programs in high-risk groups have made small HCC detectable. Partial hepatic resection (HR), liver transplantation, or local ablation treatment can all be used to treat small HCC.. (5)

Many nonsurgical ablative methods have been developed, such as Cryoablation (6), percutaneous ethanol injection (PEI) (7), acetic acid injection (8), radiofrequency ablation (RFA) (9), microwave coagulation (10), and Transcatheter arterial chemoembolization (TACE) (11). RFA is a promising and recently discovered ablation method among these medicines. It causes profound heat harm to hepatic tissue while leaving the normal parenchyma unaffected. Its basic principle involves the generation of high-frequency alternating current, which causes ionic agitation and heat conversion, followed by intracellular water evaporation, which causes irreversible cellular changes such as intracellular protein denaturation, melting of membrane lipid bilayers, and coagulative necrosis of individual tumor cells. (5)

RFA is now widely utilized as a treatment option for individuals with minor HCCs who are not candidates for HR. However, it is still debatable whether it can compete with surgery as a first-line therapy. The outcomes of published trials that looked at the effectiveness of RFA and HR for small HCC were mixed. Huang et al. (12) and Yun et al.(7) reported that HR were more favorable regardless of tumor size. Elsewhere, Chen et al. (13) and Feng et al. (14) showed that RFA was as effective as HR in the treatment of small HCCs. Additionally, Nashikawa et al. (15) and Peng et al. (16) recommended RFA as the first-line treatment for small HCCs

In Japan, the Japan Society of Hepatology (JSH) published “Evidence-based clinical guidelines for the diagnosis and treatment of HCC” in 2005, which was revised in 2009, and the “Consensus-based clinical practise manual for HCC,” which recommends: I hepatectomy for a single tumour regardless of tumour size, but local treatment may be chosen for a 2-cm or smaller tumour in Child–Pugh B patients; (ii) hepatectomy or local treatment. The American Association for the Study of Liver Disease (AASLD) supports local therapy for three or fewer 3-cm or smaller early-stage HCCs and 2-cm or smaller very-early-stage HCCs with complications such as portal hypertension across Europe and North America. RFA is advised for three or fewer 3-cm or smaller HCCs, however the standard treatment algorithms in Japan, North America, and Europe varied somewhat. (17) (18)

With the growth of technology and the need for a good quality of life, minimally invasive technology has become increasingly appealing to patients and health care professionals in recent decades, particularly in the treatment of tiny solid tumours. (19)

Radiofrequency ablation (RFA) is more effective and has fewer problems and shorter hospital stays. RFA can also be used on a regular basis. Although RFA may eventually gain acceptance as a therapy option, its long-term effectiveness and safety should be thoroughly assessed. (20) (18) Finally, due to the small number of RCTs conducted thus far, the heterogeneity of different trials, and the inherent limitations of meta-analyses, it is still uncertain whether RFA or RES (HR) is more successful for the treatment of resectable HCC patients. To compare RFA and RES(HR) treatment techniques, solid data is necessary.

## Methodology

### Data source

The text keywords “RFA versus HR” and “hepatocellular carcinoma therapy” were used in an automated search of PubMed, EMBASE, OVID, Web of Science, the Cochrane Library, Google Scholar, and the Controlled Trials Meta Register. By carefully examining the reference lists of relevant retrieved papers, more research were discovered. The sole permitted language was English. The only investigations that achieved results were those conducted by humans.

### Eligibility Criteria

The researchers searched for papers that compared RFA closure to HR closure. Research papers were excluded from the analysis along with abstracts, letters, comments, editorials, expert opinions, reviews without original data, and case reports if [1] it was impossible to extract the appropriate data from the published articles; [2] there was significant overlap between authors, institutes, or patients in the published literatures; [3] the measured outcomes were not clearly presented in the literatures; and [4] the measured outcomes were not clear.

### Study Identification

The author read all of the titles and abstracts found by the search method. Two non-author impartial reviewers independently reviewed relevant entire papers for qualifying requirements.

### Data extraction

Each qualifying paper was independently evaluated by two reviewers. Each article was analysed for the number of patients, their age, gender, use of technique, 3 year and 5 year mortality rate, duration of hospital stay and recurrence of incidence. Further discussion or consultation with the author and a third party was used to resolve conflicts. The study’s quality was assessed using the modified Jadad score.

### Statistical analysis

All of the data was obtained and entered into analytic software. Fixed- or random-effects models were used to estimate mean difference, standardised mean difference (SMD), odds ratios, and relative risk (RR) with 95 percent confidence intervals to examine critical clinical outcomes (CIs). Statistical heterogeneity was measured with the χ^2^; P < 0.100 was considered as a representation of significant difference. I^2^ greater than or equal to 50% indicated the presence of heterogeneity. Funnel plots were used to assess potential publication bias based on the prevalence of wound infection after surgery. A statistically significant difference was defined as P<0.05.

## Results

Meta-analysis showed that RFA was associated with significant decrease in the length of hospital stay for RCTs as seen in Figure 1 (SMD = -2.171, CI = -2.381 to - 1.962, p=<0.001). this results can be supported by (13) (12) (14) As depicted in Figure 2, Non-RCTs also show a significant decrease in the length of hospitalization (SMD = -1.048, CI = 1.492 to -0.937, p=<0.001) which can be supported by(21) (22) (23)

**Figure 1:**
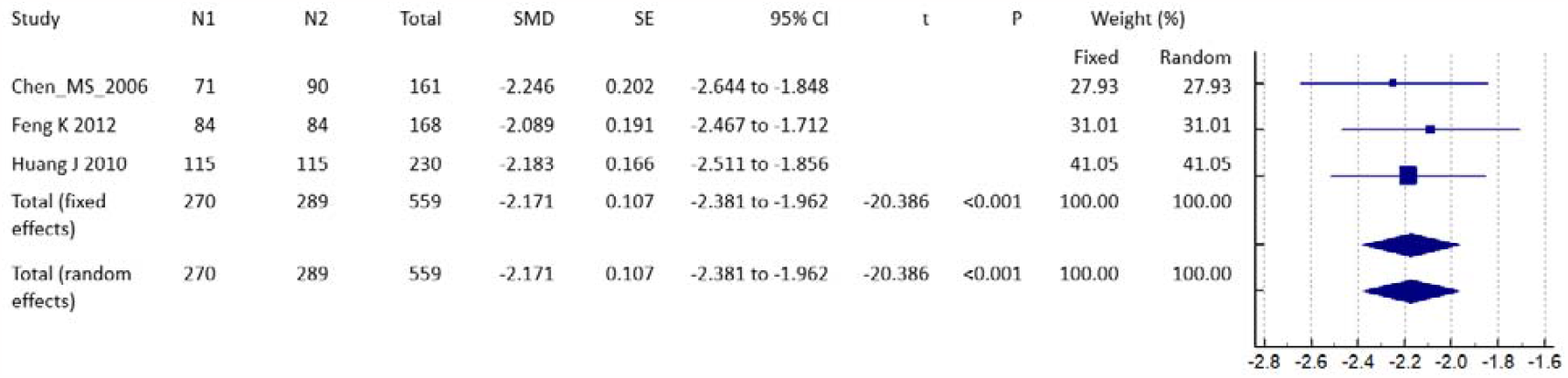
Duration of Hospital stay (RCTs)

**Figure 2:**
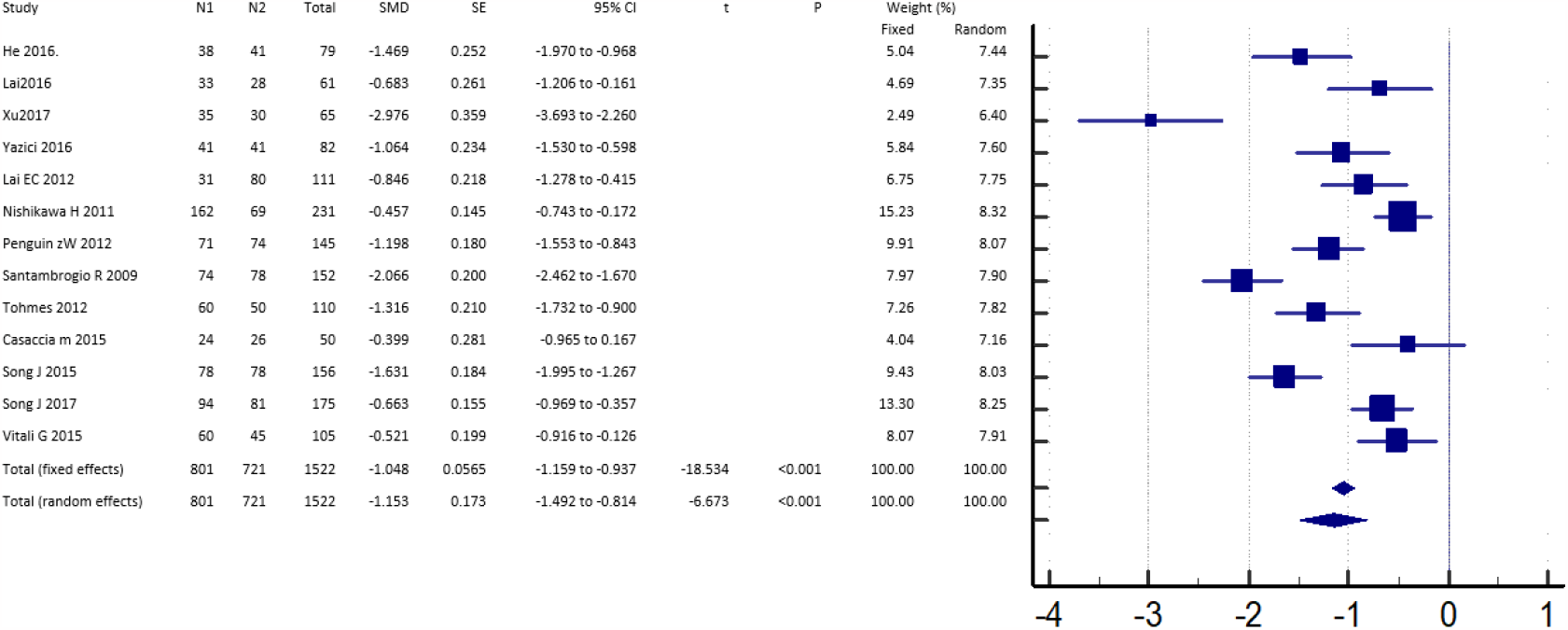
Duration of Hospital stay (Non-RCTs)

However,3-year survival chance as seen in figure 3 for RCTs (RR = 0.850, 95% CI = 0.772 to 0.935, p=0.001) and figure 4 for non-RCTs (RR = 0.941, 95%CI = 0.927 to 0.956, p=<0.001) are poorer. 5-year survivability was also poor as seen in figure 5(RR=0.856, 95% CI = 0.835 to 0.878, p=0.001). papers like (13) (14) (23) show not much difference in 3 year survivability and(24) (15)show similar 5 year survivability.

**Figure 3:**
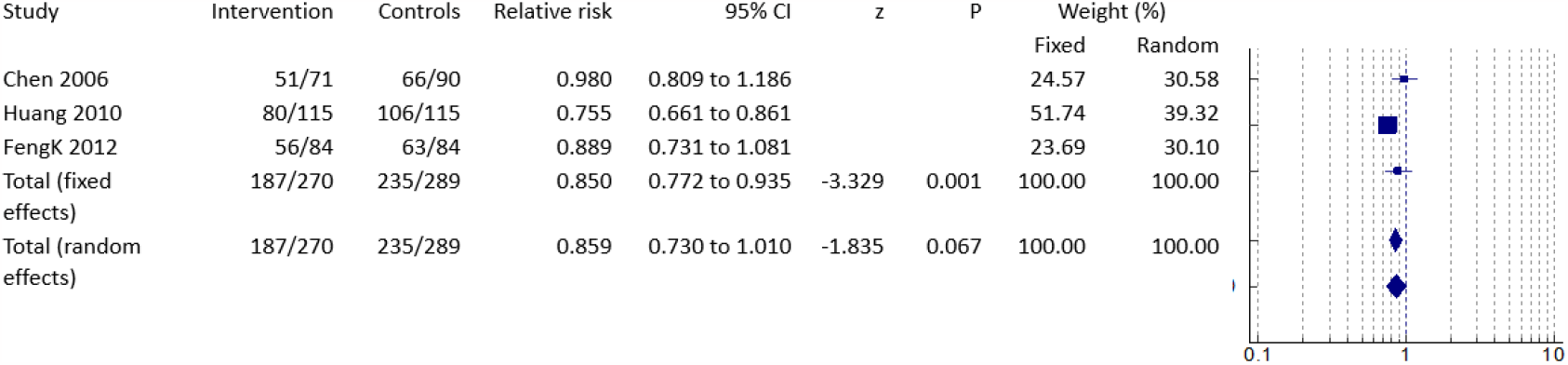
3 year survival (RCTs)

**Figure 4:**
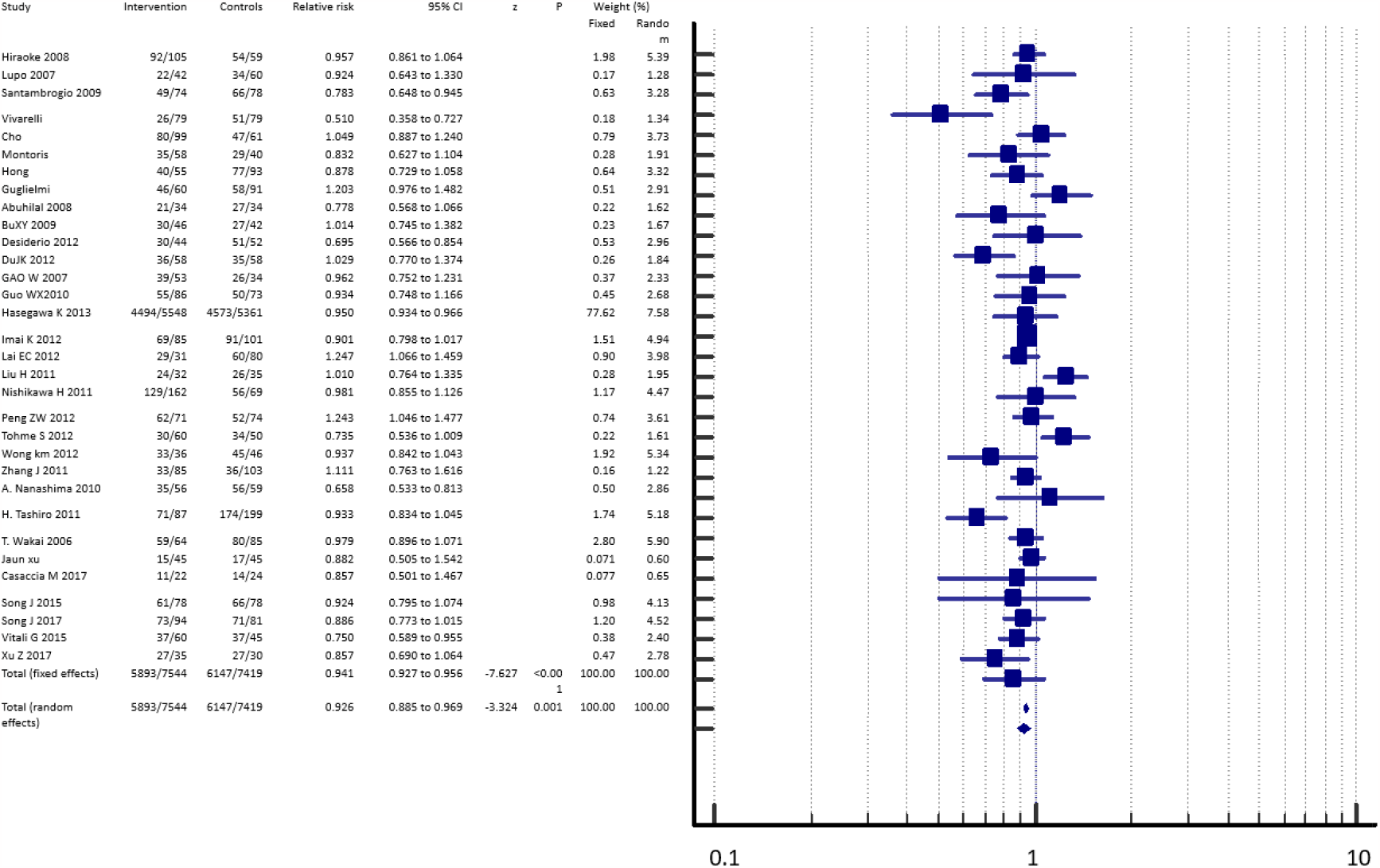
3 year survival (Non-RCTs)

**Figure 5:**
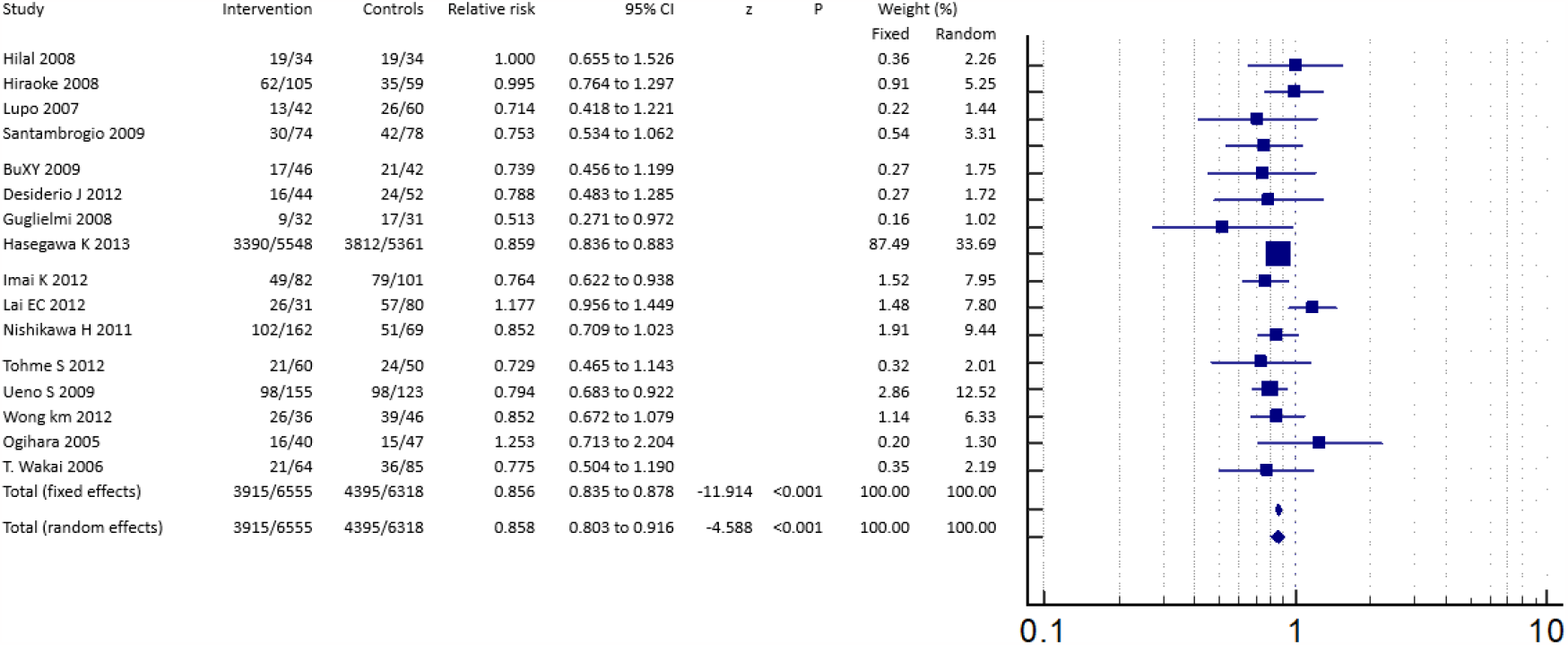
5 year survival

As it can be denoted from figure 6, it was also associated with significant increase incidence of recurrence (RR = 1.749, 95% CI = 1.444 to 2.119, p=<0.001) and more or less all studies show a higher incidence rate.

**Figure 6:**
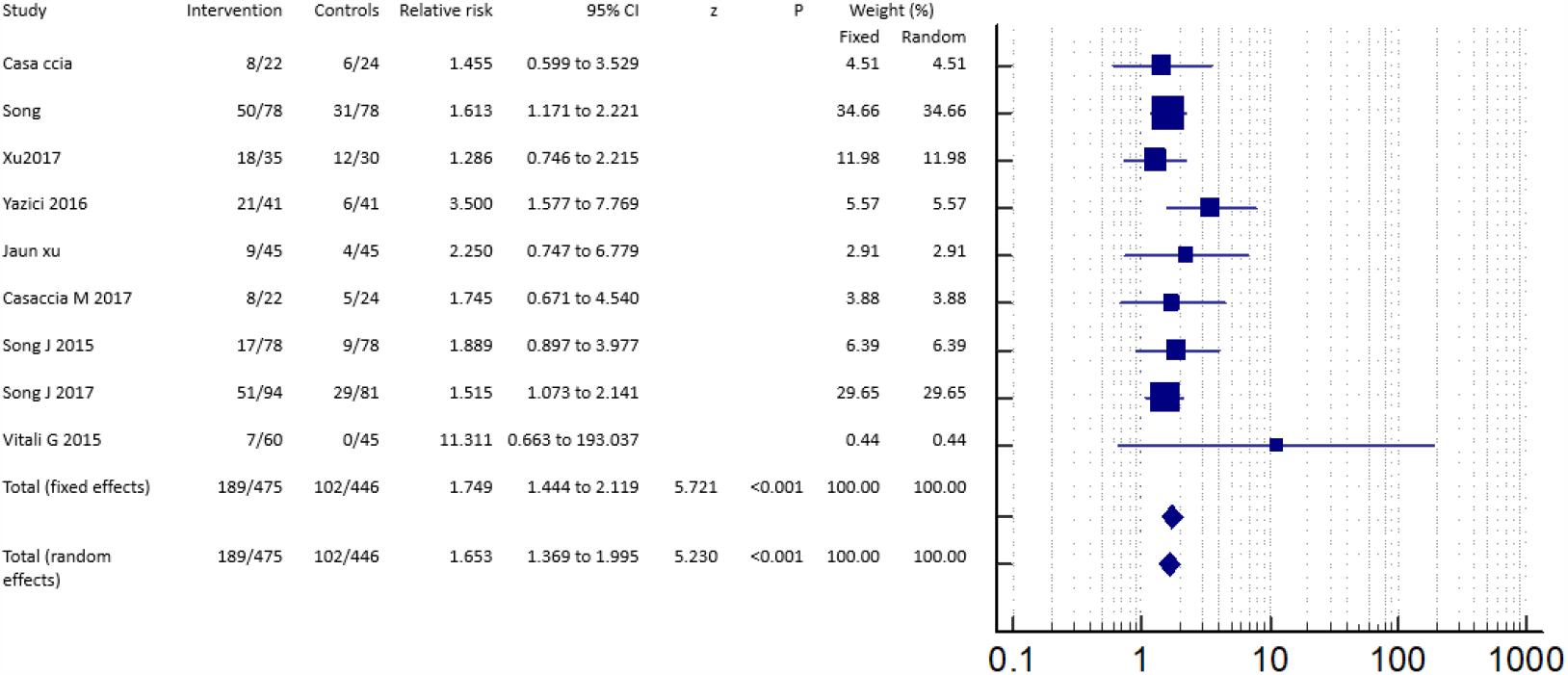
Incidence of recurrence

## Discussion

RES(HR) and RFA are now the most widely utilised and recognised as curative treatments for individuals with small HCC tumours. Under the direction of colour Doppler ultrasonography and CT, percutaneous or laparoscopic techniques for RFA therapy are often used to ablate and eliminate the lesion and surrounding tissue (25). However, there is no clear consensus on which modality is the most successful. OS and DFS are two often used key indicators for evaluating the curative impact of cancer therapy. Each index focuses on a different aspect. DFS is a major measure that reflects the therapeutic benefit of therapeutic modalities used, whereas OS represents the reaction to the entire condition, which includes complete treatment modalities, patient health, and other relevant aspects that affect survival. Despite the fact that DFS is believed to be the most appropriate measure for evaluating the effect of the treatment modalities utilised, both DFS and OS rates were used in the current study to evaluate the therapeutic effectiveness of RES and RFA. (18)

This meta-analysis reveals that HR therapy is better than RFA treatment in patients with minor HCC. (26) Surgical resection exhibited considerably superior overall survival rates at 3, and 5 years, as well as lower recurrence rates. This might be explained in part by improved surgical procedures and a better understanding of liver segmental architecture, which have resulted in a substantial drop in operative mortality and a better surgical outcome. (27) (20) Furthermore, some clinicians have struggled to comprehend the new approach and have been unable to notice modest signs of complication and recurrence, which has resulted in this outcome. Patients’ survival was further harmed by the delay in observation during successful RFA therapy. (27) (28)

The major cause of late mortality in patients with HCC is a high risk of intrahepatic recurrence following ablation therapy and/or surgical resection. Recurrence was shown to be more common after RFA than after HR in the current investigation. Inadequate ablation of the main tumour and/or the existence of tumour vascular infiltration in the neighbouring liver may cause recurrences following RFA. The original tumour and venous tumour thrombi might be removed with surgical resection. Percutaneous microwave ablation involves puncturing and burning normal tissues, which might create difficulties. The fact that surgical resection is an open technique in which defined areas with margins are removed reduces the odds of recurrence and increases survival in contrast to RFA, which is a relatively close treatment in comparison to HR, must be considered. When compared to the HR group, the RFA group exhibited greater recurrence rates at 3 and 5 years and lower complication rates. It is generally understood that tumour size, number of lesions, location, liver function, portal vein invasion, vascular invasion, and the width of the tumor-free margin following surgical excision are independent prognostic variables impacting patient survival.

Despite the greater risk of death and recurrence, our research found that RFA was linked with a shorter hospital stay than HR. RFA can be done without general anaesthesia in clinical practise. The majority of individuals getting percutaneous RFA only need to stay for 2–3 days..

However, because HCC of more than 2 cm had a greater prevalence of vascular invasion than HCC of 2 cm or less, the positive impact of HR was observed to be more significant in patients with HCC of more than 2 cm.(29) RFA is being employed as a first-line therapy option for patients with HCC tumours up to 5 cm in size (30). The therapeutic outcome is typically thought to be better the smaller the lesion. In patients with HCC tumours measuring less than 2 cm, Peng et colleagues(16) found that percutaneous RFA significantly increased OS rates but not recurrence-free survival rates when compared to RES. (18)

## Conclusion

Although RFA was linked to a shorter hospital stay, it was also linked to a higher risk of recurrence when compared to hepatic resection. The 3-year survival rate, like the 5-year survival rate, was lower. This demonstrates that HR is a more effective therapeutic option for liver cancer than RFA.

**Table 1:** Description of Papers

| Table 1 :- Description of Papers |  |  |  |  |  |  |  |  |  |  |
| --- | --- | --- | --- | --- | --- | --- | --- | --- | --- | --- |
| N<br>o. |  | Type<br>of<br>stud<br>y | Year of<br>publishi<br>ng | Time<br>period<br>of data<br>collecti<br>on | Samp<br>le<br>size | Mean<br>age | Male<br>/<br>femal<br>e | Type<br>of<br>liver<br>canc<br>er | Interventi<br>on | Quality<br>Assessm<br>ent |
| 1 | Vivarelli et al. (31) | NRCT | 2004 |  | 158 | 66.5±8.5 | 63/18 | hcc | RFA, RES | Good |
| 2 | Hong et al. (32) | NRCT | 2005 | 1999-2001 | 148 | 55.5±9.8 | 55/18 | hcc | RFA, RES | Good |
| 3 | cho et al. (33) | NRCT | 2005 | 2000-2002 | 160 | 57.5 | 62/16 | hcc | RFA, RES | Good |
| 4 | Montorsi et al. (34) | NRCT | 2005 | 1997-2003 | 98 | 67±7.5 | 42/10 | hcc | RFA, RES | Good |
| 5 | Chen et al. (13) | RCT | 2006 | 1999-2004 | 161 | 50.8 ± 11.1 | 65/15 | hcc | RFA , RES | Good |
| 6 | Lupo et al. (35) | NRCT | 2007 | 1999-2006 | 102 | 67.6 | 38/10 | hcc | RFA, RES | Good |
| 7 | Gao et al. (36) | NRCT | 2007 | 1999-2006 | 87 | 54.8 | 38/9 | hcc | RFA, RES | Good |
| 8 | Zhou et al. (37) | NRCT | 2007 | 2001-2006 | 87 | 55 ± 13.5 | 36/8 | hcc | RFA, RES | Good |
| 9 | Hiraoka et al. (38) | NRCT | 2008 |  | 164 | 65.4±9.8 | 65/ 2 | hcc | RFA, RES | Good |
| 10 | Hilal et al. (24) | NRCT | 2008 | 1991-2003 | 68 | 66 | 26/5 | hcc | RFA, RES | Good |
| 11 | Guglielmi et al. (39) | NRCT | 2008 | 1996-2006 | 200 | 65.4±9.5 | 75/20 | hcc | RFA, RES | Good |
| 12 | Abu-Hilal et al. | NRCT | 2008 | 1991-2003 | 68 | 68±6 | 26/7 | hcc | RFA, RES | Good |
| 13 | Santambrogio et al. (22) | NRCT | 2009 | 1997-2007 | 152 | 68/7 | 56/18. | hcc | RFA, RES | Good |
| 14 | Bu XY et al. (40) | NRCT | 2009 | 2000-2006 | 88 | 54.9 ± 8.8 | 38/6 | hcc | RFA, RES | Good |
| 15 | Curley et al. (41) | RCT | 2010 | 2003-2005 | 230 | 55.92±13 | 82/35 | hcc | RFA, RES | Good |
| 16 | Ueno et al. | NRC<br>T | 2010 | 2000-2005 | 278 | 66.6 | 90/4<br>8 | hcc | RFA,<br>RES | Good |
| 17 | Guo et al.<br>(42) | NRC<br>T | 2010 | 2002 -<br>2007 | 159 | 51.5 | 60/1<br>9 | hcc | RFA,<br>RES | Good |
| 18 | Yun WK et<br>al. (7) | NRC<br>T | 2010 | 2003 -<br>2007 | 470 | 53.8 ±<br>9.8 | 185/<br>52 | hcc | RFA,<br>RES | Good |
| 19 | Hung et al.<br>(43) | NRC<br>T | 2011 | 2002-2007 | 419 | 64.1 ±<br>11.8 | 162/<br>58 | hcc | RFA,<br>RES | Good |
| 20 | Liu et al.<br>(44) | NRC<br>T | 2011 | 2008-2010 | 67 | 33.5 | 27/6 | hcc | RFA,<br>RES | Good |
| 21 | Nishikawa<br>et al. (15) | NRC<br>T | 2011 | 2004-2010 | 231 | 68.2 ±<br>9.2 | 75/3<br>8 | hcc | RFA,<br>RES | Good |
| 22 | Wang et al.<br>(45) | NRC<br>T | 2011 | 2002-2009 | 595 |  | 215/<br>66 | hcc | RFA,<br>RES | Good |
| 23 | Zhang et<br>al. (46) | NRC<br>T | 2011 | 2006-2009 | 188 | 57.5 ±<br>13.5 | 78/<br>15 | hcc | RFA,<br>RES | Good |
| 24 | Feng et al.<br>(14) | RCT | 2012 | 2005-2008 | 168 | 49 | 77/6 | hcc | RFA,<br>RES | Good |
| 25 | Du JK et<br>al. (47) | NRC<br>T | 2012 | 2003-2007 | 116 | 57.5±8.<br>4 |  | hcc | RFA,<br>RES | Good |
| 26 | Song et al.<br>(23) | NRC<br>T | 2015 |  | 156 | 48 | 70/8 | hcc | RFA,<br>RES | Good |
| 27 | He et al.<br>(48) | NRC<br>T | 2016 |  | 79 | 53.8±<br>10.5 | 29/5 | hcc | RFA,<br>RES | Good |
| 28 | Lai et al.<br>(49) | NRC<br>T | 2016 |  | 61 | 59.2±<br>11.8 | 22/2<br>6 | hcc | RFA,<br>RES | Good |
| 29 | Yazici et<br>al. (50) | NRC<br>T | 2016 |  | 82 | 7 2.8±<br>5.2 | 24/1<br>7 | hcc | RFA,<br>RES | Good |
| 30 | Casaccia<br>et al. (51) | NRC<br>T | 2017 |  | 46 | 62.6±8.<br>5 | 17/6 | hcc | RFA,<br>RES | Good |
| 31 | Wang et al.<br>(52) | NRC<br>T | 2017 |  | 126 | 65.<br>8±15.3 | 35/2<br>8 | hcc | RFA,<br>RES | Good |
| 32 | Xu et al.<br>(21) | NRC<br>T | 2017 |  | 65 | 55.6±10<br>.8 | 26/6 | hcc | RFA,<br>RES | Good |

## Data Availability

All data produced in the present work are contained in the manuscript

